# The rise of SARS-CoV-2 variant B.1.1.7 in Israel intensifies the role of surveillance and vaccination in elderly

**DOI:** 10.1101/2021.02.16.21251819

**Authors:** A Munitz, M Yechezkel, Y Dickstein, D. Yamin, M Gerlic

**Author notes:** Corresponding authors: Dan Yamin, Faculty of Engineering, Tel Aviv University Ramat Aviv 69978, Israel., Ariel Munitz and Motti Gerlic PhD, The Faculty of Medicine, Tel-Aviv University, Ramat Aviv 69978, Israel., respectively. These authors contributed equally (A Munitz, D. Yamin, M Gerlic).

## Abstract

Since the emergence of the SARS-CoV-2 pandemic various generic variants have been described. Of specific interest is a new variant, which was observed in England during December 2020 and is now termed B.1.1.7. This variant is now associated with increased infectivity and therefore its spread within the community is of great importance. The Israeli government established three noteworthy programs namely, mass PCR testing, focused protection of the elderly and more recently an unparalleled prioritized vaccination program. In this study we analyzed primary data of >300,000 RT-PCR samples collected throughout December 6th 2020 until February 10th 2021 in the general community and nursing homes. We identified that within a period of six weeks, the B.1.1.7 variant was capable of out competing the wildtype SARS-CoV-2 strain to become the main strain. Furthermore, we show that the transmission of B.1.1.7 in the 60+ population reached a near complete halt, due to an ongoing surveillance testing program in nursing homes and the vaccination program of Israel. Thus, proactive protection programs such as routine surveillance and monitoring of populations at risk combined with prioritized vaccination, is achievable and will result in a reduction of severe illness and subsequent death.

## Introduction

In December 2020, a distinct phylogenetically cluster of SARS-CoV-2 was identified in London as well as in Southeast and East of England^1^. Since the emergence of this variant (now termed variant B.1.1.7), it has been sequenced and has shown to display a number of mutations. These mutations may affect its infectivity and thus spread in the community^2^. Variant B.1.1.7 is defined by 17 mutations, among which several are located in the spike protein that mediates SARS-CoV-2 attachment and entry into human epithelial cells^3^. At least two mutations were suggested to have biological significance. Mutation N501Y has been shown to enhance binding affinity to human angiotensin converting enzyme 2 (ACE2)^3^, while mutation P681H, was suggested to affect infection and transmission due to its location^4,5^. In support of this, preliminary evidence from epidemiological studies estimated that B.1.1.7 is 43-82% more transmissible^6^ and thus associated with an increase in the effective reproduction number ***R***_***t***_ by a factor of 1.4-1.8^7^.

During the COVID-19 pandemic, Israel has established three noteworthy programs. The first, is a high throughput national RT-PCR testing program that is based on large laboratories, capable of assessing up to 9,200 tests/million inhabitants per day. The second, an ongoing surveillance testing program in nursing homes (also termed “Protector of Fathers and Mothers” program) (https://corona.health.gov.il/en/magen-israel/). Thirdly, an unparalleled pro-active national vaccination program (using the Pfizer vaccine), that reached a coverage of 80% for the first dose in elderly >60 within 38 days^8^. To explore the transmission dynamics of the variant B.1.1.7 and to estimate the success of the above operations to mitigate the risk in the general population and in the elderly, we analyzed primary data of >300,000 RT-PCR samples collected throughout December 6^th^ 2020 until February 10^th^ 2021 in the general community and nursing homes. This timely data provides an ideal setting to shed light on future policy making of governments in light of the possibility spread of the B.1.1.7 variant or even future ones.

## Results

### Rapid transmission of the B.1.1.7 variant in the Israeli Population

The B.1.1.7 variant poses a deletion of two amino acids at positions 69-70 (Δ69-70), which is associated with the inability of the Thermo Fisher TaqPath COVID-19 assay probe to detect the Spike gene (S gene). Indeed, S gene target failures (SGTF) were recently shown to be primarily due to the new variant^9^, and molecular studies have used this “Flat S” phenomenon to assess the transmission data of the B.1.1.7 variant^9^. Using SGTF data from available ∼300,000 individual tests, which were analyzed throughout December 6^th^ 2020 until February 10^th^ 2021 we were able to monitor the spread of B.1.1.7 variant and assess the impact of Israel programs to reduce transmission. Until December 22, the B.1.1.7 variant was undetectable within the pool of positive cases in Israel. Strikingly, within a period of four weeks, the B.1.1.7 variant became the dominant strain (January 21^st^), reaching to 92% following two more weeks (Figure 1A). Specifically, we calculated the **effective reproduction number, *R***_***t***_ which is the expected number of new infections at time *t* caused by an infectious individual. To evaluate the daily *R*_*t*_, we used the methodology previously introduced by Cori et al.^10,11^ that was also considered in our previous work^12,13^. This data-driven approach suggests that *R*_*t*_ is the ratio between *I*_*t*_, the number of new infections generated at day t and 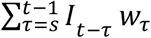, the sum of infection incidence up to time step t - 1 weighted by the infectivity function *w*_*τ*_. For *w*_*τ*_, we considered 10 days truncated gamma distribution with a mean of 3.96 days (95% CI: 3.53-4.39 days), an SD 4.75 days (4.46-5.07 days)^14^. We used our data to evaluate the reproductive number for both the SARS-COV-2 wild type and the B.1.1.7 variant. For an accurate representation of Israel, we scaled our data with the age-stratified incidence data published daily by the Israeli ministry of health (MOH) (https://datadashboard.health.gov.il/COVID-19/). Finally, we shifted the data by 3.5 days, which corresponded to the average time between exposure until obtaining the test outcome. Prior the impact of the national lockdown the *R*_*t*_ of the B.1.1.7 variant was as high as 1.71 (95% CI: 1.59-1.85) compared to 1.12 (95% CI: 1.10-1.15) observed for the wildtype (Figure 1B, December 30, 2021). Furthermore, we found that during the entire period the variant was 45% more transmissive then the wildtype.

**Figure 1.**
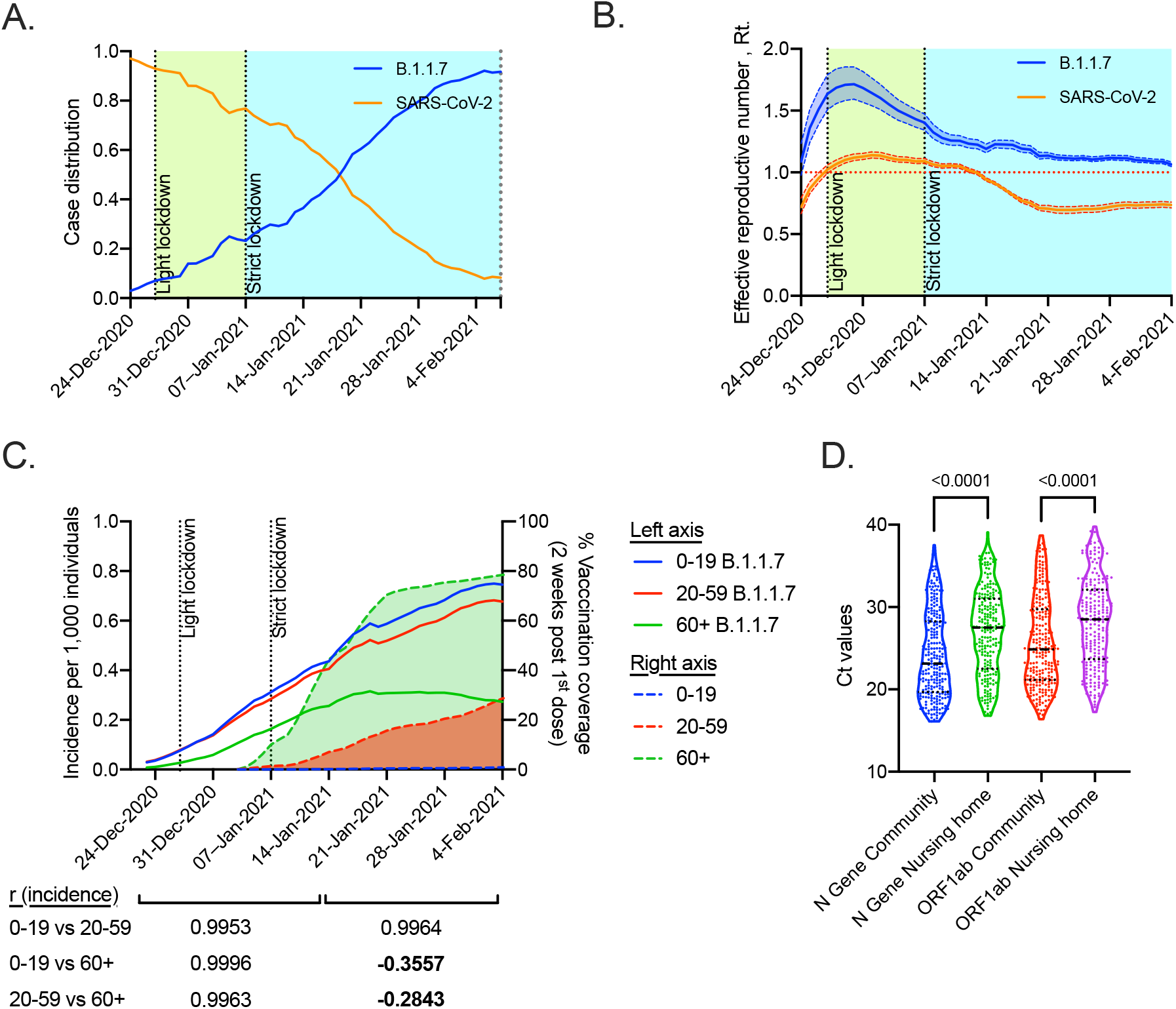
The emergence of variant B.1.1.7 in Israel. (A) Daily new cases distribution between SARS-CoV-2 and variant B.1.1.7 over time. (B) Effective reproduction number of SARS-CoV-2 and variant B.1.1.7 with 95% credible intervals over time. The effective reproduction number, R_t, was calculated based on the methodology previously introduced by Cori et al. (C) Incidence per 1,000 individuals of the variant B.1.1.7 stratified by age (left axis). Cumulative vaccination coverage two weeks after 1st dose per age group (right axis). Data were calculated using GraphPad Prism 9; Correlation analysis was performed using a Pearson correlation coefficient test (two-tailed, 95% confidence). r values are shown. (D) The Ct values distribution (presented as a violin plot) for N gene and ORF1ab gene among infected individuals above 60 years at nursing homes versus infected individuals above 60 years at the general community. Data were calculated using GraphPad Prism 9; the black dotted line represents the calculated median and quartiles values. Statistical analysis was performed using a t-test, P values are shown.

Assessment of the distribution of the B.1.1.7 variant in different age groups revealed a clear increase in the numbers of B.1.1.7^positive^ individuals from age groups 0-19 and 20-59. While a modest increase was observed in the 60+ aged group until January 14, 2021, the incidence of B.1.1.7^positive^ individuals aged 60+ plateaued and eventually declined (Figure 1C). In support of this, pearson correlation analysis of the rise in incidence among the different age groups revealed that between December 24, 2020 and January 14, 2021 the rise in the incidence of B.1.1.7 was highly correlated among all age groups (r>0.99, Figure 1C). Following January 14, 2021, a striking decline was observed in the correlation coefficient only in the 60+ age group (r= −0.35 and −0.28, 60+ vs. 0-19 or 20-59 age, respectively, Figure 1C). Notably, this phenomenon was associated with the fact that by that time point, 50% of the 60+ aged population was two weeks post the 1^st^ dose of vaccination.

To assess the ongoing surveillance testing program in nursing homes, we compared the Ct threshold values in 60+ age groups in the general community versus nursing homes. Ct threshold values were used as predictive markers for viral load where a higher Ct indicates lower mucosal viral titers. This analysis revealed significantly lower Ct values, in both viral tested genes, in the community in comparison with nursing homes with matching age groups (t-test, *p<0*.*001*) (Figure 1D). Collectively, as viral load drives transmission^15,16^ these data underscores the important role of random surveillance testing in nursing homes.

## Discussion

Using data collected through RT-PCR testing at the Electra-TAU lab we investigated the dynamics and spread of the B.1.1.7 variant. We identified that within a period of six weeks, the B.1.1.7 variant was capable of out competing the wildtype SARS-CoV-2 strain to become the main strain (more than 90% of positive tests). We found that the B.1.1.7 variant is 45% more transmissible than the wildtype strain (95% CI: 20-60%). This is within the range of previous estimates, which suggested a 1.56 increase in England^9^. These similar estimates were observed despite the substantial differences between the countries in terms of demographic characteristics including age distribution, household size, the density of living areas, as well as key settings that impact transmission including lockdowns and vaccination coverage.

Increased viral transmission may have substantial implications on the ability to protect SARS-CoV-2 susceptible populations. Thus, it is important to examine governmental policies such as vaccination programs and focused protection interventions. Comparative age-based analysis demonstrated that transmission and spread of the B.1.1.7 variant among the majority of the population at risk (as define by being over the age of 60) was significantly reduced in comparison with other age groups. Of even greater interest, we show that while the transmission of B.1.1.7 continued to rise dramatically in the population aged 0-59 with similar kinetics, the rise in the 60+ population reached a near complete halt. This is likely due to impact of the Israeli vaccination program, which was initiated in the 60+ age group. Consequently, a clear containment of B.1.1.7 variant was shown in 60+ age group after January 14. In this regard, by this date in which the B.1.1.7 variant already gained transmission dominance, ∼50% of the elderly population were two weeks post their 1^st^ vaccination dose. This interpretation is highly reasonable given the relative rise in B.1.1.7 transmission in ages 0-59 during that same time and the fact that no other measurements were introduced.

Our data confirmed that pro-active surveillance programs of populations at risk such as those found in nursing homes were capable of early detection, which likely enabled containment of further viral spread within this housing community. This is observed by the significant difference in Ct threshold levels, which were higher in nursing homes in comparison with the general population. Thus, proactive protection programs such as routine surveillance and monitoring of populations at risk combined with prioritized vaccination, is achievable and will result in a reduction of severe illness and subsequent death.

## Data Availability

All data are available upon request from the authors.

## Ethics

This study was approved by the Tel Aviv University Institutional review board (IRB# 0002746-2).

## Author contributions

MA, DY and GM, collected the data; MA, YM, YD and GM design and analyzed the data; MA, YM, YD and GM wrote and edited the manuscript; MA, YD and GM supervised the work.

